# Automatic classification of seizure adequacy in electroconvulsive therapy: A proof-of-concept study

**DOI:** 10.64898/2026.11.11.26367803

**Authors:** Majda Panah, Benedicte Lumby Jessen, Amalie Rand, Tejs Jansen, Martin Balslev Jorgensen, Morten Mørup, Anders Jorgensen

**Author notes:** Correspondence: Anders Jorgensen MD PhD DMSci, Psychiatric Center Copenhagen, Bispebjerg-Frederiksberg Hospital Nordre Fasanvej 57, DK-2000 Frederiksberg, Denmark. Joint first authors (Majda Panah, Benedicte Lumby Jessen). Joint last authors (Morten Mørup, Anders Jorgensen).

## Abstract

Electroconvulsive therapy (ECT) is the most effective treatment of a range of severe mental disorders and is annually given to more than a million patients worldwide. Dosing of the electrical charge is critical for efficacy and side-effect burden; however, it is currently based on manual assessment of seizure quality and exhibits substantial clinical heterogeneity. Here, we collected 1,000 ECT session electroencephalograms (EEG) from 130 individual ECT patients and made an expert-level classification of the EEGs into actionable seizure quality categories (null-response [0], threshold [T], and adequate [A]). We then trained a range of machine learning algorithms on the raw EEG data to classify the EEGs into these categories and tested for classification accuracy. The highest performing algorithm was able to classify the seizures with an area under the receiver operating characteristic curves of >0.9 for both the 0 vs. T and the T vs. A categories corresponding to excellent discrimination. In contrast, the accuracy of predicting seizure quality in subsequent treatments was only moderate. The addition of clinical variables to the models, such as demographics, anesthesia and anticonvulsant exposure, did not improve model performance. These results provide proof-of-concept of machine learning classification of seizure adequacy in ECT based on raw EEG data. The findings could pave the way for the development of more objective and personalized decision-support tools in ECT.

## Introduction

Electroconvulsive therapy (ECT) remains the most acutely effective treatment for a range of prevalent and severe psychiatric disorders, including depression, mania and mixed states, and catatonia ^1–5^. It also holds efficacy in treatment resistant schizophrenia ^6^. ECT is estimated to be given to approximately 1.4 million patients worldwide every year ^7^, translating into an estimated total of 15 million individual ECT sessions. Consequently, ensuring the delivery of high-quality ECT is a key clinical concern in psychiatry.

ECT relies on the induction of a generalized seizure by applying a small electrical current through the brain tissue^5^. The seizure is recorded in an electroencephalogram (EEG). Based on whether the EEG classification is deemed adequate (above seizure threshold) or inadequate (at or below seizure threshold), the charge is either maintained or increased at the next treatment. Misclassification of a seizure as adequate can lead to ineffective treatments and a prolonged time to recovery. Conversely, misclassification of seizures as inadequate can lead to an unnecessary increase of the charge, thereby worsening cognitive side effects ^8,9^. No single EEG characteristic is sufficient in deeming seizures as adequate or inadequate, neither do they individually show strong association to treatment efficacy ^10^. Hence, determination of the charge is based on an overall assessment of the seizure architecture, taking into account EEG characteristics such as amplitude, coherence, and post-ictal suppression of the seizure ^10–12^.

Currently, assessing the seizure adequacy and adjusting the electrical charge relies entirely on the manual judgement of the ECT clinician. These clinicians – either medical doctors or nurses – often possess varying levels of expertise and experience in ECT ^13^. Furthermore, although it is well established that seizure threshold can increase during a course of ECT ^14,15^, reliable predictors of subsequent seizure adequacy remain unavailable. Consequently, dosing is largely based on a trial-and-error approach, where the dose is adjusted only after an inadequate seizure occurs.

An observational study of 3,343 patients comprising a total of 41,909 ECT procedures across five Danish psychiatric departments revealed profound institutional variations in ECT practices ^14^. We found an up to two-fold difference in age-adjusted electrical charge at initiation, a four-fold difference by the tenth treatment, and a more than 3-fold difference in the number of reported inadequate seizures ^14^. Such extreme variation is likely to substantially influence patient-level treatment outcomes, including time to recovery and cognitive side-effects. Similar variation presumably exists across other countries and healthcare systems.

The use of machine learning is increasingly considered a path forward to enable data-driven precision psychiatry, and ECT is an area of particular interest in this regard ^16,17^. Most studies applying machine learning methodologies in ECT have been focusing on prediction of treatment outcome from clinical and structural magnetic resonance imaging data, finding moderate prediction accuracy ^18–20^. To our knowledge, only two previous studies have used machine learning for EEG characterization in ECT, one focusing on separating ictal from non-ictal EEG segments and reproduce ECT quality indices calculated by the stimulation device ^21^, and another using expert-rated seizure adequacy indices to obtain an overall adequacy score ^22^.

Here, our primary aim was to use machine learning to analyze the raw, unprocessed EEG data recorded during ECT to classify the seizure into clinically relevant, actionable seizure adequacy categories. Our secondary aim was to predict the seizure adequacy category at the subsequent treatment.

## Subjects and Methods

### Data collection and study cohort

The study utilized two linked data sources: structured clinical treatment records extracted from the electronic health record system (Epic) and raw EEG recordings acquired during individual ECT-sessions. Each observation was defined per individual ECT session and thus every patients contributed with multiple observations across a treatment course. EEG data were collected from ECT procedures performed at Psychiatric Center Copenhagen within the Capital Region of Denmark, where approximately 3,000 ECTs are performed annually. The current digital EEG collection system was implemented during 2025, using a freely available software provided by the ECT device manufacturer ^23^. The present analyses included treatments performed from November 1^st^, 2025, to April 1^st^, 2026, comprising over 1,000 EEGs from 130 individual ECT patients. EEG collection completeness was >90%.

The structured clinical dataset contained information on patient demographics, treatment characteristics, medication exposure, seizure duration, motor seizure duration, electrode placement, stimulation dose, anesthesia and the seizure adequacy label assigned during routine clinical care. EEG recordings from Psychiatric Center Copenhagen were acquired during ECT treatments using the Thymatron System IV device set to the 2X dose program, which allows for a charge increase up to 200% (1008 mC). The standard electrode placement is bitemporal, which is utilized in approximately 90% of patients ^14^. Right unilateral placement could be selected at the first or subsequent treatments, if clinically indicated. Treatments are given by medical doctors (3 times per week) or nurses (once per week).

The device records two-channel EEG activity throughout the induced seizure from the time of the electrical stimulation. For each treatment, the Thymatron device generates a PDF report containing both a visual representation of the EEG trace and an embedded comma separated values file with the raw EEG signal. These raw signal files formed the basis for EEG preprocessing and feature extraction.

Clinical records and EEG recordings were using unique patient identifiers and treatment dates. Exclusions were applied to duplicate clinical records, duplicate EEG recordings, unmatched EEG files, recordings with uncertain linkage, and treatments with insufficient EEG adequacy for reliable interpretation.

### EEG classification

To ensure an expert-level classification of the EEGs on which the machine learning algorithm was trained, a physician with special experience and training in EEG interpretation in ECT (MP) evaluated all the EEGs. We based the seizure classification on a recent and authoritative systematic review on the ictal EEG in ECT ^10^. Here, key individual seizure characteristics emphasized to be associated with clinical effect was recruitment phase duration, midictal amplitude, interhemispheric coherence, and postictal suppression. In borderline cases, peak heart rate was also taken into consideration ^24,25^. Based on these key parameters, overall seizure adequacy was classified as one of 4 categories: Adequate (Type A), Threshold (Type T), overdosed (Type S), and no seizure (Type 0). Type S only occurred in <5 instances and was left out of the analyses. All artifacts and their duration were also recorded. All seizures not immediately classifiable as type A was further evaluated by 2 senior ECT psychiatrists (AJ and MBJ), and a consensus decision was made. The expert reviews were blinded to the original clinician-assigned labels during relabeling.

### EEG data preprocessing and feature extraction

Raw EEG recordings were processed using a standardized Python-based pipeline. Signal values were converted to numeric format and transformed into MNE Raw objects ^26^ to allow reproducible EEG processing. Because the Thymatron recordings were stored in microvolts and MNE expects signals in volts, all signals were converted accordingly. To reduce noise and remove irrelevant frequency components, a zero-phase 50 Hz FIR notch filter was applied to attenuate power line interference, followed by a zero-phase 0.5–45 Hz FIR band-pass filter.

Since ECT-induced seizures evolve over time, features were extracted both from the full EEG recording and from predefined temporal segments. Global features were intended to summarize the overall seizure morphology, whereas segment-based features were included to capture changes between the early, middle, and late parts of the seizure, such as the build-up of ictal activity and subsequent suppression. Each recording was segmented into overlapping two-second epochs with 50% overlap, and power spectral density was estimated using Welch’s method.

The final EEG representation included spectral, temporal, amplitude, coherence, and suppression-related features. Spectral features comprised absolute and relative band power across conventional EEG frequency ranges, including delta, theta, alpha, and beta bands. To summarize the distribution of spectral power over time, percentile-based statistics were computed across epochs. In addition, band power was calculated within predefined temporal segments to capture seizure evolution across the recording. Broader spectral shape was represented using spectral centroid, peak frequency, and spectral entropy.

Amplitude features were estimated from the signal envelope using the Hilbert transform and summarized both globally and within temporal segments. Inter-channel synchrony was quantified using magnitude-squared coherence between the two EEG channels, both across the full recording and within frequency bands. Suppression-related features were derived from differences between high- and low-power periods and were included to reflect postictal suppression dynamics. In total, 200 EEG-derived features were extracted for each treatment session (for a full description of feature engineering, see Supplementary Material).

In addition to EEG-derived features, selected clinical and treatment variables known to potentially influence seizure architecture were considered. This included sex, age at treatment, electrode placement, treatment number within series, stimulation dose, cumulative benzodiazepine exposure, and cumulative antiepileptic drug exposure ^15,27,28^. Anesthesia-related variables included total administered doses of thiopental, propofol, and etomidate. Because anesthesia data were incomplete, models including anesthesia features were evaluated only in the subset of observations with complete anesthesia information.

### Training of machine learning algorithms for seizure classification

Two related modelling tasks were investigated. The first modelling task was classification of the observed seizure adequacy at the current ECT session using EEG-derived features, with or without additional clinical and anesthesia-related variables. Models were trained both as a three-class problem, distinguishing 0, T, and A seizures, and as a binary problem restricted to T vs. A seizures. Several supervised learning models were evaluated, including a linear ordinal probit model, two ordinal probit artificial neural network models (OPANN-1 and OPANN-2), a decision tree classifier, and an extreme gradient boosting classifier (XGBoost). The ordinal probit models were based on the cumulative ordinal regression framework proposed by Chu and Ghahramani ^29^ in which an underlying continuous latent score is mapped to the ordered seizure adequacy categories through learned thresholds. The ordinal models were included because seizure adequacy has a natural clinical ordering (0 < T < A), allowing this prior information to be incorporated directly into the model structure. In contrast, the decision tree and XGBoost models were included as non-ordinal benchmark models capable of capturing non-linear relationships and feature interactions without explicitly modelling the ordinal structure of the outcome.

All ordinal probit models were implemented in PyTorch using a shared custom framework based on the cumulative ordinal regression model ^29^. The linear ordinal probit model consisted of a single fully connected linear layer producing a latent score. In OPANN-1 and OPANN-2, the linear predictor was replaced by feed-forward neural networks with two hidden layers of 32 and 16 neurons and 64 and 32 neurons, respectively. All hidden layers used rectified linear unit (ReLU) activation functions and dropout regularization (dropout = 0.2). The output layer consisted of a single neuron representing the latent score, which was transformed into class probabilities using the ordinal probit likelihood. The predicted seizure adequacy class was defined as the class with the highest predicted probability.

The decision tree classifier was implemented using the DecisionTreeClassifier class from scikit-learn with the Gini impurity criterion. The gradient boosting model was implemented using the XGBClassifier class from the XGBoost library. Both models were trained to estimate the probability of each seizure adequacy class, and the final prediction was defined as the class with the highest estimated probability.

The machine learning methodology was designed to prioritize robust internal validation. Model development and evaluation were performed using 10 repeated five-fold nested cross-validation runs, in which the outer loop was used to estimate model performance and the inner loop was used for hyperparameter optimization. Because several patients contributed multiple treatment sessions, all cross-validation splits were grouped by patient to ensure that observations from the same patient never appeared in both training and test data, thereby preventing patient-level information leakage and providing a more realistic estimate of generalization to previously unseen patients. Stratification was used to approximately preserve the distribution of seizure adequacy labels across folds.

The ordinal probit models were trained by minimizing the negative log-likelihood of the observed ordinal outcomes using the Adam optimizer with L2 regularization implemented through weight decay. Hyperparameters for all models were selected within the inner cross-validation loop. For the ordinal probit models, the learning rate and weight decay were optimized. For the decision tree classifier, the maximum tree depth and minimum number of samples per leaf were optimized, whereas for XGBoost the learning rate and maximum tree depth were optimized. Model performance was primarily evaluated across the 10 repeated runs using different patient-level data partitions. In addition, the linear ordinal probit model was evaluated using fixed patient-level data partitions and repeated random parameter initializations to isolate the variability attributable to model initialization. Models were also evaluated with and without class weighting, where applicable, to reduce the influence of class imbalance. Class weights were calculated inversely proportional to the class frequencies in the training data within each fold and applied only during model fitting. Throughout the Results section, Probit refers to the linear ordinal probit model, whereas OPANN-1 and OPANN-2 refer to the two neural-network-based ordinal probit models. For the Probit model, Data Splits denotes experiments performed using different patient-level cross-validation partitions across the repeated runs, whereas Initialization denotes experiments performed using fixed patient-level data partitions and different random model initializations.

Performance was evaluated using the macro-averaged F1-score, which gives equal weight to each class and is therefore appropriate for imbalanced classification tasks, balanced accuracy, and the receiver operating characteristic area under the curve (ROC-AUC). In the three-class setting, pairwise ROC-AUC values were calculated for 0 vs. T and T vs. A seizures, whereas in the binary setting ROC-AUC was calculated for T vs. A seizures. For the ordinal models, ROC-AUC was calculated from the continuous latent seizure adequacy score, whereas for the decision tree and XGBoost models it was calculated from the predicted class probabilities.

For each repeated cross-validation run, predictions from the outer test folds were pooled before calculating the performance metrics, resulting in a single performance estimate per run. Results are reported as mean ± standard deviation across the 10 repeated runs. To contextualize model performance against routine clinical assessment, the expert consensus labels were compared with the original clinician-assigned labels using confusion matrices, overall accuracy, balanced accuracy, and Cohen’s kappa.

### Training of machine learning algorithms for seizure adequacy prediction

The second modelling task was prediction of seizure adequacy at the subsequent ECT session. This analysis was designed to explore whether information from a previous treatment session, together with the planned change in stimulation dose, could predict whether the next treatment would result in an adequate seizure.

For this analysis, consecutive treatment pairs were constructed within each patient. Treatment *t* was paired with treatment *t*+1 only when the treatments were consecutive within the same treatment series. The prediction target was the expert-labelled seizure adequacy at treatment *t*+1. Input features included EEG-derived characteristics from treatment t, the previous seizure label, treatment number, stimulation dose at treatment t, and the planned dose difference between treatment *t* and treatment *t*+1.

This resulted in 733 paired observations for the three-class prediction task, with target labels comprising 654 type A seizures, 72 type T seizures, and seven type 0 seizures. A binary T vs. A prediction dataset was also constructed, comprising 719 paired observations with 648 type A and 71 type T target labels. For the subsequent-outcome prediction task, a reduced set of models were evaluated, consisting of the linear ordinal probit model, the larger ordinal neural network, decision tree, and XGBoost. The same patient-grouped nested cross-validation framework, class weighting strategy, and evaluation metrics were used as described for the classification task.

Finally, a post hoc dose-response analysis was conducted for the binary subsequent-outcome prediction task. For each test observation, the hypothetical stimulation dose at treatment *t*+1 was varied across a predefined range while holding all other patient and treatment features fixed. Predicted probabilities of an A seizure were then estimated across the dose range. This analysis was performed to examine whether the models captured a clinically plausible relationship between stimulation dose and predicted seizure adequacy. Because dose values were varied post hoc within an observational dataset, this analysis was interpreted as a model-based sensitivity analysis rather than a causal estimate of the effect of stimulation dose.

### Ethics

The extraction and use of the data for research purposes was approved by the Capital Region of Denmark (protocol number R-24030479) in accordance with the Danish Health Care legislation (*Sundhedsloven* §46). Individual informed consent is waived for this type of study.

## Results

### Characterization of the study cohort

The final classification dataset consisted of 961 valid treatment sessions from 125 unique patients. Of these 75 (60%) were female and 50 (40%) were male. The mean age was 51.7 ± 19.6 years (range 18–90). The most prevalent primary diagnosis was unipolar depression (48.0%), followed by schizophrenia (24.0%), bipolar depression (8.0%), and mania (4.8%). 14.4% of patients had other primary diagnoses. The mean number of ECT treatments per patient was 7.7 ± 5.0 (range 1–26).

### ECT practitioner vs. expert classification of seizures

To contextualize model performance relative to routine clinical assessment, expert consensus labels were compared with the seizure adequacy labels assigned by clinicians during routine ECT care. Overall agreement was high, with an accuracy of 0.93. However, agreement was more moderate after accounting for class imbalance and chance agreement, as reflected by a Cohen’s kappa of 0.547. Balanced accuracy was therefore more informative than overall accuracy and reached 0.846 in the three-class setting and 0.902 when restricted to the binary T-versus-A distinction (Figure 1).

**Figure 1:**
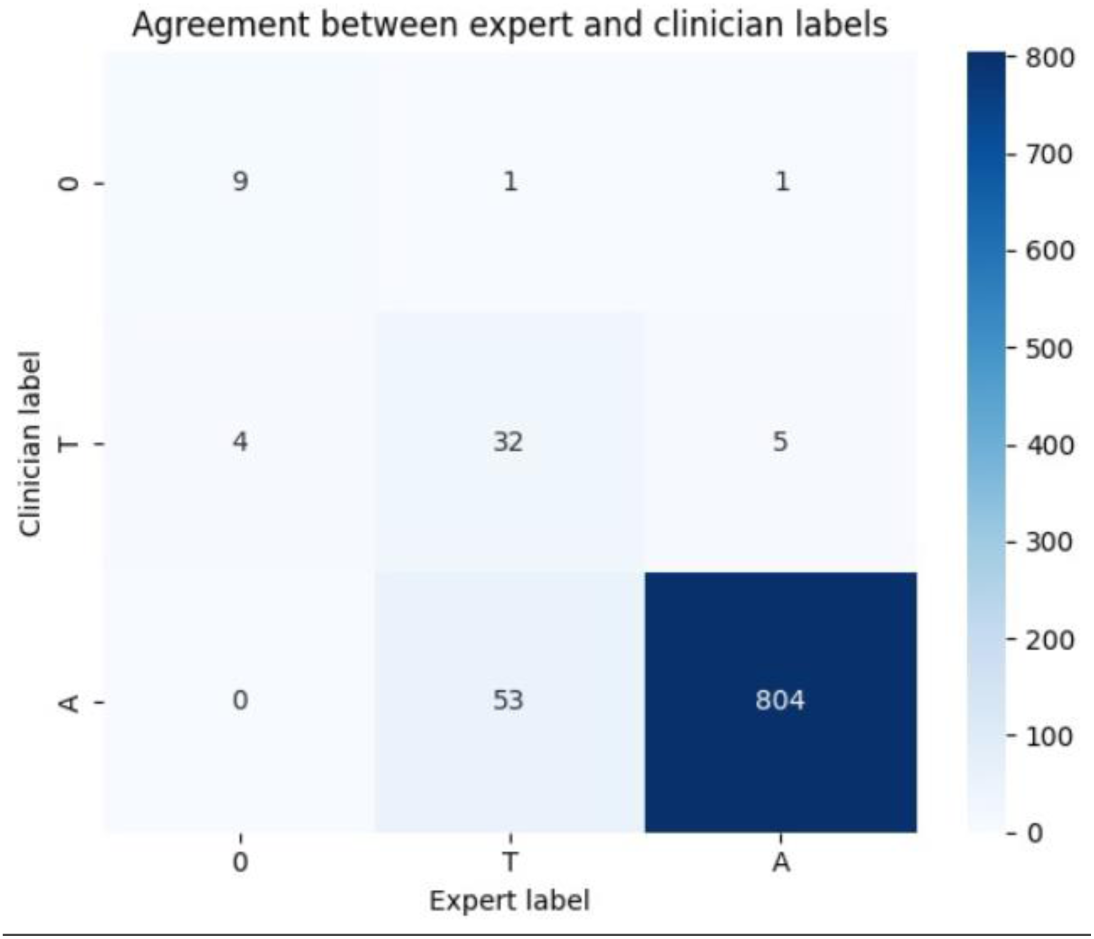
Confusion matrix illustrating disagreement patterns between clinician and expert seizure classifications.

The confusion matrix showed that disagreement occurred primarily between neighboring seizure categories, particularly T and A seizures. This distinction is clinically important because it directly informs the subsequent stimulation strategy. The expert-clinician comparison therefore provides a relevant clinical benchmark for interpreting the machine learning results, where model performance should be considered in the context of the variability already present in routine EEG-based seizure assessment.

### Seizure classification performance of machine learning algorithms

Classification of observed seizure adequacy showed the strongest overall model performance. Across models and configurations, discrimination between T and A seizures was consistently high. ROC-AUC values for this distinction were generally above 0.85 across all model types, indicating that EEG-derived features contained clinically meaningful information for separating seizures that would usually lead to dose escalation from those where the current dose would usually be maintained. In contrast, discrimination between 0 seizures and T seizures was more variable, reflecting the very small number of no-seizure observations and the greater difficulty of modelling this rare class (Table 1).

**Table 1:** Receiver operating characteristic area under the curve (ROC-AUC) performance for all machine learning models models in classification of expert-labelled seizure adequacy in electroencephalograms (EEGs) recorded from 961 electroconvulsive therapy sessions. Data are showed for analyses with vs. without weighting of class imbalance and with vs. without the inclusion of clinical variables into the models (see text for details).

| **Model** | **Configuration** | **AUC (O vs T)** | **AUC (T vs A)** |
|---|---|---|---|
| Linear Probit (Data Splits) | EEG, Weights | 0.903 ± 0.035 | 0.904 ± 0.009 |
|  | EEG, No Weights | 0.691 ± 0.014 | 0.929 ± 0.006 |
|  | EEG+Clin., Weights | 0.889 ± 0.018 | 0.902 ± 0.008 |
|  | EEG+Clin., No Weights | 0.703 ± 0.053 | 0.915 ± 0.007 |
| Linear Probit (Initialization) | EEG, Weights | 0.891 ± 0.044 | 0.902 ± 0.010 |
|  | EEG, No Weights | 0.680 ± 0.013 | 0.932 ± 0.003 |
|  | EEG+Clin., Weights | 0.840 ± 0.047 | 0.900 ± 0.011 |
|  | EEG+Clin., No Weights | 0.712 ± 0.026 | 0.917 ± 0.009 |
| OPANN-1 | EEG, Weights | 0.850 ± 0.071 | 0.907 ± 0.014 |
|  | EEG, No Weights | 0.342 ± 0.069 | 0.913 ± 0.019 |
|  | EEG+Clin., Weights | 0.805 ± 0.091 | 0.903 ± 0.016 |
|  | EEG+Clin., No Weights | 0.337 ± 0.081 | 0.925 ± 0.007 |
| OPANN-2 | EEG, Weights | 0.902 ± 0.079 | 0.924 ± 0.016 |
|  | EEG, No Weights | 0.303 ± 0.051 | 0.926 ± 0.003 |
|  | EEG+Clin., Weights | 0.850 ± 0.098 | 0.913 ± 0.008 |
|  | EEG+Clin., No Weights | 0.322 ± 0.061 | 0.925 ± 0.011 |
| Decision Tree | EEG, Weights | 0.608 ± 0.086 | 0.852 ± 0.024 |
|  | EEG, No Weights | 0.677 ± 0.062 | 0.853 ± 0.029 |
| XGBoost | EEG, Weights | 0.842 ± 0.023 | 0.939 ± 0.022 |
|  | EEG, No Weights | 0.671 ± 0.019 | 0.960 ± 0.002 |

In the three-class setting, the weighted EEG-only ordinal probit model achieved ROC-AUC values of 0.903 for 0 seizure versus T seizure and 0.904 for T versus A seizure. The corresponding ROC curves illustrate that the model learned a continuous seizure-adequacy score with clinically meaningful separability between adjacent seizure categories (Figure 2).

**Figure 2:**
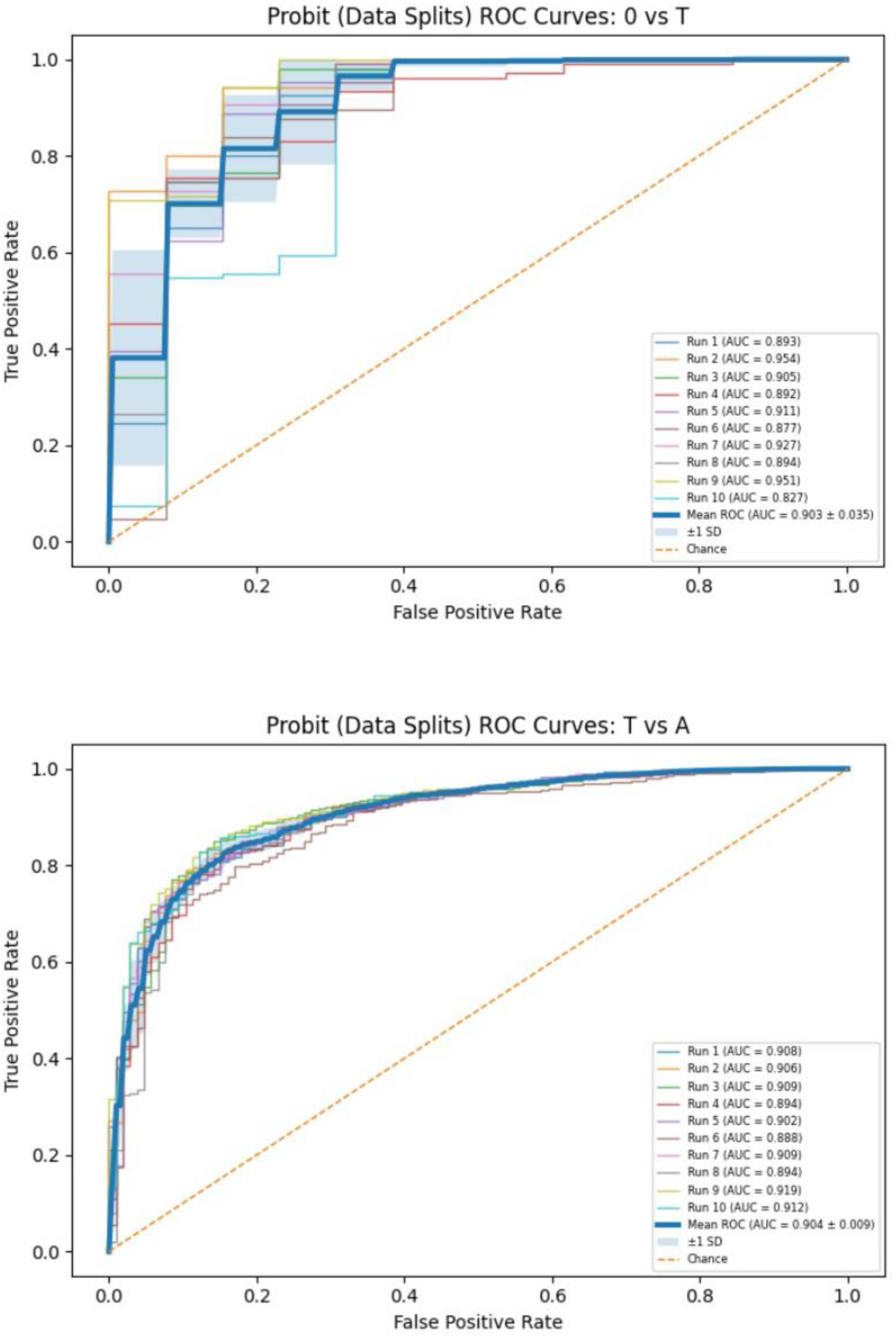
Receiver operating characteristic (ROC) curves for the weighted EEG-only ordinal probit model. Top: no seizure vs threshold seizure. Bottom: Threshold seizure vs adequate seizure.

The best overall classification performance was achieved by XGBoost trained on EEG-derived features with class weighting, with a macro F1-score of 0.792 and balanced accuracy of 0.837. The linear ordinal probit model also performed well in the weighted EEG-only configuration, with a macro F1-score of 0.665 and balanced accuracy of 0.809. Although the ordinal neural network models were included to allow more flexible non-linear relationships, they did not outperform the simpler linear ordinal probit model in the three-class setting. This suggests that, given the available sample size and feature representation, additional neural network flexibility did not translate into improved generalization. Full three-class classification performance across model configurations is provided in Table S1.

Class weighting improved performance particularly for the ordinal models. This was most evident in balanced accuracy, indicating that weighting increased sensitivity to the underrepresented seizure categories rather than merely improving overall accuracy. Without class weighting, models tended to favor the majority class, A seizures, resulting in poorer recall for T-and 0-seizure observations. The effect of class weighting was less pronounced for XGBoost and the decision tree.

Adding structured clinical variables did not improve classification performance beyond EEG-derived features alone. Across the ordinal models, inclusion of these clinical variables led to negligible or slightly reduced performance. Similarly, anesthesia-related variables did not provide systematic improvement in the subset of observations with complete anesthesia information. These findings suggest that clinically relevant information contained in these variables may already be reflected in the EEG recordings, or that the available structured variables did not provide additional independent discriminatory information for expert-labelled seizure adequacy.

In the binary T-versus-A classification setting, performance improved across models compared with the three-class task. This supports the interpretation that much of the difficulty in the three-class setting was driven by the rare no-seizure class. The binary task also represents the most clinically central distinction, as it corresponds to whether the stimulation dose should usually be maintained or increased at the next treatment. Across models, ROC-AUC values remained high, further supporting that EEG-derived features can distinguish T from A seizures. Full binary classification performance across models is provided in Table S2.

### Predictive performance of machine learning algorithms

Prediction of seizure adequacy at the subsequent ECT session was substantially more difficult than classification of the observed seizure. In the three-class prediction setting, all models achieved only modest performance. Macro F1-scores remained below 0.40, and balanced accuracy was only slightly above the three-class baseline. This indicates that future seizure adequacy could not be predicted reliably at the individual level from the available previous-treatment information. Full three-class subsequent-outcome prediction performance is provided in Table S3.

Because no-seizure outcomes were very rare, and because the distinction between T and A seizures is most relevant for dose planning, a binary prediction task was also evaluated. In this setting, performance improved but remained moderate (Table 2). Hence, in the binary prediction task, XGBoost achieved the highest macro F1-score and ROC-AUC, with a macro F1-score of 0.601 and AUC of 0.722. The linear ordinal probit model achieved the highest balanced accuracy, with a mean of 0.663. Nevertheless, all models remained only moderately above baseline, indicating that prediction of the next seizure outcome is substantially more challenging than classification of the current seizure.

**Table 2:** Binary prediction performance of all machine learning models for subsequent seizure outcome in electroencephalograms (EEG) recorded from electroconvulsive therapy sessions.

| **Model** | **F1-score** | **Balanced Accuracy** | **AUC (T vs A)** |
|---|---|---|---|
| Linear Probit | 0.5723 ± 0.0194 | 0.6626 ± 0.0272 | 0.7090 ± 0.0260 |
| OPANN-2 | 0.5329 ± 0.0334 | 0.6521 ± 0.0401 | 0.6826 ± 0.0635 |
| Decision Tree | 0.5566 ± 0.0184 | 0.6080 ± 0.0295 | 0.6161 ± 0.0320 |
| XGBoost | 0.6011 ± 0.0143 | 0.6534 ± 0.0240 | 0.7223 ± 0.0318 |

### Dose-response analysis

To examine whether the prediction models captured clinically plausible relationships between stimulation dose and subsequent seizure adequacy, a post hoc dose-response analysis was performed. For each test observation, the hypothetical stimulation dose at the next treatment was varied while all other features were held fixed. This analysis should be interpreted as a model-based sensitivity analysis, not as a causal estimate of the effect of dose.

Most models produced largely flat dose-response curves, indicating that varying the hypothetical dose had little effect on predicted seizure outcome probabilities. The main exception was the binary linear ordinal probit model, which showed a smooth monotonic increase in the predicted probability of an A seizure as the hypothetical dose increased. This suggests that the binary probit model captured a population-level dose-response pattern that was not apparent in the more flexible models.

Age-stratified dose-response curves from the binary probit model suggested a lower average predicted probability of an EEG-defined A seizure among patients aged 60 years or older compared with younger patients across the examined dose range (Figure 3).

**Figure 3:**
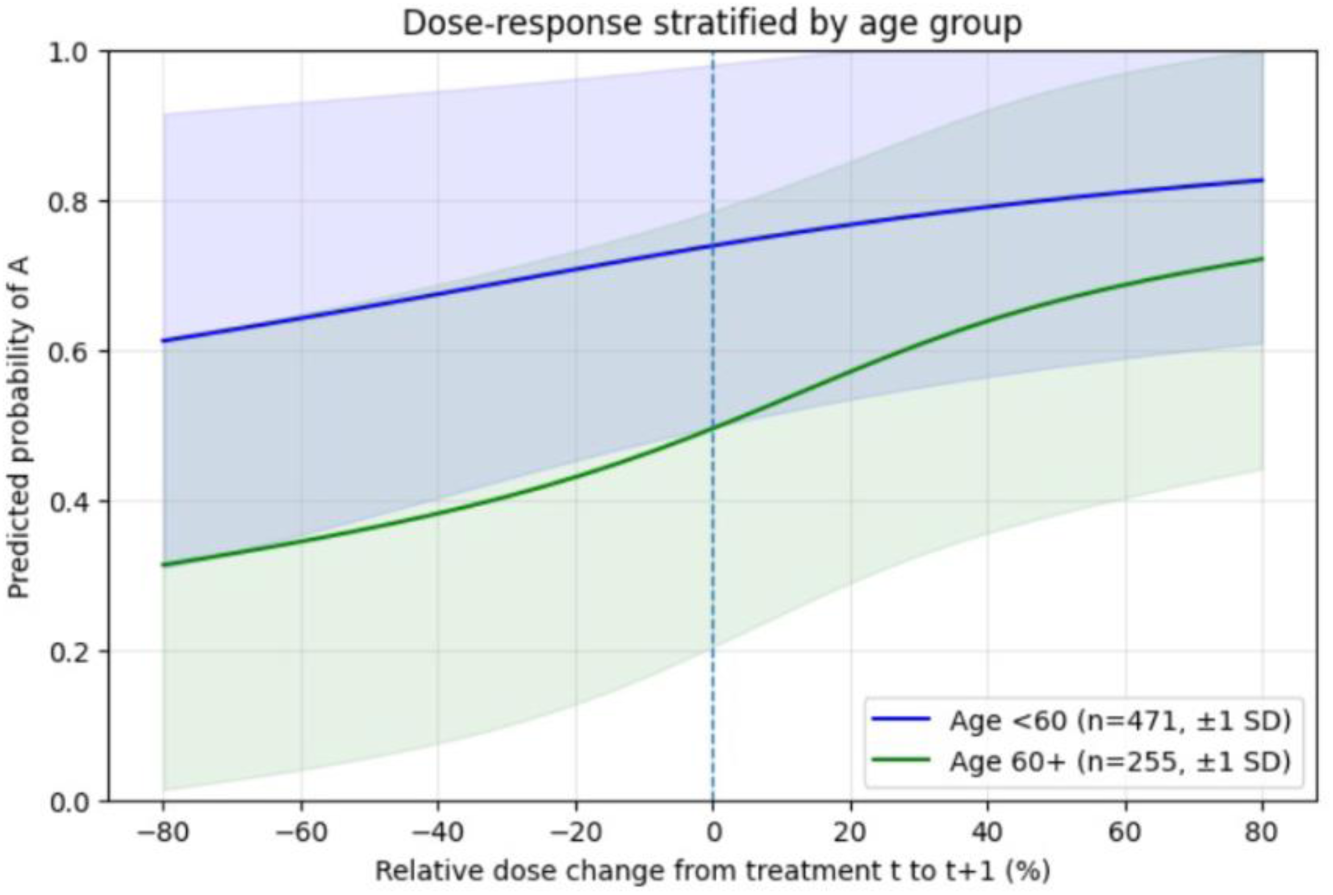
Dose response in the binary setting averaged across observations and stratified by age, computed for a single run.

Although the mean predicted probability was lower in the older group, there was substantial variability within both age groups and considerable overlap between patient-specific curves. The age-stratified pattern should therefore be interpreted as a population-level tendency rather than a reliable individual-level prediction. One possible explanation is that EEG-defined seizure adequacy may vary with age-related physiological characteristics, meaning that older patients may be less likely to show EEG patterns classified as adequate even when stimulation dose is increased. Importantly, this does not necessarily imply reduced therapeutic benefit, as the model target was EEG-defined seizure adequacy rather than clinical antidepressant response.

## Discussion

In this study, we present a proof of concept that machine learning algorithms can be applied to accurately classify seizure adequacy in ECT from raw EEG data. Based on a dataset of 1,000 EEGs, we obtained high ROC-AUC values for the discrimination between null-response, threshold, and adequate seizures. The algorithms were trained on EEGs labeled for seizure adequacy by experienced ECT clinicians and based on known seizure quality variables. The inclusion of contextual information on ECT-technical and sociodemographic variables did not improve model performance. Taken together, these findings indicate that machine learning-based algorithms can act as a clinically valuable translator of raw EEG data into actionable seizure adequacy categories, which can in turn directly guide the ECT-clinician regarding the strategy for the subsequent treatment. These could include seizure-improving initiatives such as lowering anesthetic dose, change anesthetic or increase the charge. The fact that the addition of sociodemographic and clinical variables did not improve model performance makes a clinical implementation of the models even more feasible, because such data are not necessarily easily obtained or incorporated outside of research settings. As such, we believe these findings could pave the way for machine learning-powered evidence-based decision support in ECT.

While machine learning has been used extensively to classify EEG segments in epilepsy and other neurological disorders, we are only aware of two previous studies using machine learning to classify seizure adequacy in ECT. In a recent study by Kayser et al ^21^, machine learning was successfully applied to separate ictal from non-ictal segments, identify the seizure endpoint and reproduce seizure quality indices calculated by the ECT device. Hence, the major difference to our study is that in the study by Kayser et al, they did not classify EEGs into overall adequacy categories, which can directly guide the dosing strategy. In a study by Al-Kaysi et al ^22^, machine learning was used to classify EEGs into adequacy categories, but the model input was manually rated seizure quality indices. Hence, the major difference to our study is that Al-Kaysi et al did not use raw EEGs as model input, which is a necessity if the model is to assist ECT dosing for ECT-clinicians in a more objective and scalable manner without the requirement of human resources and interaction. Hence, we consider our findings to significantly contribute to machine learning applications in ECT and potentially move us forward towards a more objective and consistent dosing of the charge.

Our secondary goal was to predict the adequacy category of the subsequent treatment. this task proved to be considerably more difficult and only yielded moderate accuracy in a binary prediction model. The difference between current-seizure classification and subsequent-outcome prediction is clinically and methodologically important. In the classification task, EEG features describe the seizure being classified. In contrast, the prediction task requires the model to infer the next seizure outcome from information available at the previous treatment, together with the planned dose change. The weaker performance therefore likely reflects both the indirect nature of the prediction target and the influence of patient-specific physiology, treatment adaptation, and clinical decision-making between sessions. Finally, we cannot rule out that the number of EEGs combined with the class imbalance impaired our ability to predict subsequent seizures.

The major advantages in our study are that it was based on raw EEG data, which can easily be recorded in every ECT clinic, to categorize the seizures into adequacy categories that can directly be used to guide clinical actions. We tested a range of machine learning algorithms using state of the art methodology, applied expert labeling of the seizures before training, and reported a range of complementary performance metrics. Finally, we had access to extensive contextual clinical information from the electronic health records. Limitations include that we did not perform an external validation on data from another department; that we only included EEGs recorded by a Thymatron device and thus cannot necessarily extrapolate the findings to settings using other devices such as the Mecta, and finally that our sample size may have been too low to provide a sufficient training set for the machine learning algorithms to achieve performance predictions. Future directions will thus include expansion of the dataset to increase precision in particular for the rarer seizure categories; testing the algorithm performance on EEG data from a different department to provide external validation, and ultimately to perform a clinical trial of the effects of the algorithm/decision support tool, which should include patient-relevant ECT outcomes such as dosage, number of treatment in the series, number of inadequate treatments in the series, as well as clinical effect and side-effects of ECT.

In conclusion, we provide proof-of-concept evidence that machine learning can be applied to classify EEGs from ECT into actionable categories of seizure categories with a high level of accuracy. A preliminary attempt to predict seizure category of subsequent treatment did not reach sufficient accuracy to be clinically implementable at this stage. Our results could lay the foundation for machine learning-driven decision support in ECT.

## Data Availability

Data from this study cannot be made publicly available

## Acknowledgements

The study was supported by the Mental Health Services of the Capital Region of Denmark and an unrestricted grant from BETA:HEALTH (grant number 2025-1985).

## References

1 James SL, Abate D, Abate KH, Abay SM, Abbafati C, Abbasi N et al. Global, regional, and national incidence, prevalence, and years lived with disability for 354 diseases and injuries for 195 countries and territories, 1990–2017: a systematic analysis for the Global Burden of Disease Study 2017. The Lancet 2018; 392: 1789–1858.

2 Mutz J, Vipulananthan V, Carter B, Hurlemann R, Fu CHY, Young AH. Comparative efficacy and acceptability of non-surgical brain stimulation for the acute treatment of major depressive episodes in adults: systematic review and network meta-analysis. BMJ 2019;: l1079.

3 UK ECT Review Group. Efficacy and safety of electroconvulsive therapy in depressive disorders: a systematic review and meta-analysis. Lancet 2003; 361: 799–808.

4 Kellner CH, Obbels J, Sienaert P. When to consider electroconvulsive therapy (ECT). Acta Psychiatr Scand 2020; 141: 304–315.

5 Espinoza RT, Kellner CH. Electroconvulsive Therapy. N Engl J Med 2022; 386: 667–672.

6 Sinclair DJ, Zhao S, Qi F, Nyakyoma K, Kwong JS, Adams CE. Electroconvulsive therapy for treatment-resistant schizophrenia. Cochrane Database Syst Rev 2019; 3: CD011847.

7 Țăpoi C, Alexander L, de Filippis R, Agorastos A, Almeida D, Bhatia G et al. Early career psychiatrists’ perceptions of and training experience in electroconvulsive therapy: A cross-sectional survey across Europe. Eur Psychiatry 2025; 67: e86.

8 Sackeim HA, Prudic J, Devanand DP, Kiersky JE, Fitzsimons L, Moody BJ et al. Effects of Stimulus Intensity and Electrode Placement on the Efficacy and Cognitive Effects of Electroconvulsive Therapy. N Engl J Med 1993; 328: 839–846.

9 Deng Z-D, Argyelan M, Miller J, Quinn DK, Lloyd M, Jones TR et al. Electroconvulsive therapy, electric field, neuroplasticity, and clinical outcomes. Mol Psychiatry 2022; 27: 1676–1682.

10 Francis-Taylor R, Ophel G, Martin D, Loo C. The ictal EEG in ECT: A systematic review of the relationships between ictal features, ECT technique, seizure threshold and outcomes. Brain Stimul 2020; 13: 1644–1654.

11 Kranaster L, Aksay SS, Bumb JM, Hoyer C, Jennen-Steinmetz C, Sartorius A. A novel Seizure Quality Index based on ictal parameters for optimizing clinical decision making in electroconvulsive therapy. Part 1: development. Eur Arch Psychiatry Clin Neurosci 2018; 268: 819–830.

12 Kranaster L, Jennen-Steinmetz C, Sartorius A. A novel seizure quality index based on ictal parameters for optimizing clinical decision-making in electroconvulsive therapy. Part 2: Validation. Eur Arch Psychiatry Clin Neurosci 2019; 269: 859–865.

13 Bjørnshauge D, Hjerrild S, Videbech P. Electroconvulsive Therapy Practice in the Kingdom of Denmark: A Nationwide Register-and Questionnaire-Based Study. J ECT 2019; 35: 258–263.

14 Jorgensen A, Andersen AK, Nielsen J, Videbech P, Jorgensen MB. Heterogeneity of Electroconvulsive Therapy Practice Under a Common Guideline: Results From a Large Electronic Health Record-Based Data Set. The Journal of ECT 2025. doi:10.1097/YCT.0000000000001194.

15 Luccarelli J, McCoy TH, Seiner SJ, Henry ME. Changes in seizure duration during acute course electroconvulsive therapy. Brain Stimul 2021; 14: 941–946.

16 Chekroud AM, Bondar J, Delgadillo J, Doherty G, Wasil A, Fokkema M et al. The promise of machine learning in predicting treatment outcomes in psychiatry. World Psychiatry 2021; 20: 154–170.

17 Lundin RM, Falcao VP, Kannangara S, Eakin CW, Abdar M, O’Neill J et al. Machine Learning in Electroconvulsive Therapy: A Systematic Review. J ECT 2024; 40: 245–253.

18 Semple DM, Suveges S, Steele JD. Electroconvulsive therapy response and remission in moderate to severe depressive illness: a decade of national Scottish data. Br J Psychiatry 2024; 225: 547–555.

19 Redlich R, Opel N, Grotegerd D, Dohm K, Zaremba D, Burger C et al. Prediction of Individual Response to Electroconvulsive Therapy via Machine Learning on Structural Magnetic Resonance Imaging Data. JAMA psychiatry 2016; 73: 557–64.

20 Bruin WB, Oltedal L, Bartsch H, Abbott C, Argyelan M, Barbour T et al. Development and validation of a multimodal neuroimaging biomarker for electroconvulsive therapy outcome in depression: a multicenter machine learning analysis. Psychol Med 2024; 54: 495–506.

21 Kayser M, Hurlemann R, Philipsen A, Freundlieb N, Kiebs M. Machine learning based seizure classification and digital biosignal analysis of ECT seizures. Sci Rep 2025; 15: 6409.

22 Al-Kaysi AM, Al-Ani A, Galvez V, Loo CK, Ling S, Boonstra TW. Estimating The Quality of Electroconvulsive Therapy Induced Seizures Using Decision Tree and Fuzzy Inference System Classifiers. In: 2018 40th Annual International Conference of the IEEE Engineering in Medicine and Biology Society (EMBC). IEEE: Honolulu, HI, 2018, pp 3677–3680.

23 GPD. https://github.com/elektrika-inc/GPD-wiki/wiki.

24 Jorgensen A, Christensen SJ, Jensen AEK, Olsen NV, Jorgensen MB. The influence of the anesthesia-to-stimulation time interval on seizure quality parameters in electroconvulsive therapy. Journal of affective disorders 2018; 231: 41–43.

25 Galvez V, Hadzi-Pavlovic D, Wark H, Harper S, Leyden J, Loo CK. The Anaesthetic-ECT Time Interval in Electroconvulsive Therapy Practice--Is It Time to Time? Brain stimulation 2016; 9: 72–7.

26 Larson E, Gramfort A, Engemann DA, Leppakangas J, Brodbeck C, Jas M et al. MNE-Python. 2026. doi:10.5281/zenodo.19666955.

27 Gillving C, Ekman CJ, Hammar Å, Landén M, Lundberg J, Movahed Rad P et al. Seizure Duration and Electroconvulsive Therapy in Major Depressive Disorder. JAMA Netw Open 2024; 7: e2422738.

28 McDonnell K, Jelovac A, Mohan C, Whooley E, Igoe A, McCaffrey C et al. Association between concomitant anticonvulsants and clinical and cognitive outcomes of electroconvulsive therapy for depression. Brain Stimul 2025; 18: 1600–1607.

29 Chu W, Ghahramani Z. Gaussian Processes for Ordinal Regression. Journal of Machine Learning Research 2005; 6: 1019–1041.

30 1.10. Decision Trees. scikit-learn. https://scikit-learn/stable/modules/tree.html (accessed 13 July2026).

