## Supplemental Material for "Automatic classification of seizure adequacy in electroconvulsive therapy: A proof-of-concept study"

Panah et al, 2026

Supplementary Material

### Feature Engineering

#### EEG Features

#### Preprocessing

Each CSV file contained raw EEG data recorded on two channels (EEG1, EEG2) during the ECT treatment. Non-numeric values were converted to numeric format to ensure continuous signals prior to further processing. The signals were then converted into an MNE Raw object to enable standardized EEG processing. The original recordings were stored in microvolts (μV), while the MNE framework expects signals in volts (V). Therefore, all values were converted from microvolts to volts by multiplying by $10^{-6.}$

EEG signals were preprocessed to reduce noise and remove irrelevant frequency components using the MNE library. A notch filter at 50 Hz was applied to remove power line interference originating from the electrical grid, using a finite impulse response filter (FIR). Subsequently a band-pass filter between 0.5 and 45 Hz was applied to remove slow signal drifts and high-frequency noise. The EEG signals were segmented into short, overlapping epochs to account for their non-stationary nature and to enable analysis of temporal dynamics and variability throughout each recording. An epoch length of 2 seconds with 50% overlap was chosen to provide sufficient temporal resolution to capture rapid changes in the EEG signal, while still allowing for reliable spectral estimation within each epoch. Since recording durations varied between 15 and 140 seconds, the number of epochs differed across recordings.

#### Spectral and Temporal EEG Features

The present feature extraction strategy was designed to capture complementary aspects of seizure activity across both time and frequency domains. Consequently, EEG representations were derived using power spectral density estimation, band-specific power measures, temporal segmentation, spectral shape descriptors and temporal dynamics features.

#### Estimation of Power Spectral Density

For each 2-second epoch (400 samples at 200 Hz), the power spectral density (PSD) was estimated using Welch's method as implemented in MNE's low-level function psd_array_welch. Each epoch was segmented into overlapping 1-second windows (200 samples) with 50% overlap, resulting in three Welch windows per epoch. The spectra estimated from these windows were averaged to obtain one PSD estimate for each epoch and EEG channel. This approach reduces the variance of spectral estimates while preserving frequency resolution and maintaining sensitivity to temporal changes in the EEG signal. The PSD was computed over the frequency range 1-30 Hz, covering the primary EEG frequency bands relevant for seizure analysis.

#### Global Band Power Features

From the PSD, total spectral power across 1-30 Hz and band-specific power were computed by integrating the PSD within predefined frequency ranges. The frequency bands comprised total power (1-30 Hz), delta (1-4 Hz), theta (4-8 Hz), low alpha (8-10 Hz), high alpha (10-13 Hz), low beta (13-20 Hz) and high beta (20-30 Hz). These bands were selected to capture physiologically distinct components of seizure-related EEG activity while providing a compact representation of the spectral distribution of the signal. For each epoch, band power values were averaged across the two EEG channels and subsequently log-transformed to stabilize variance and reduce the influence of extreme values.

To characterize the distribution and temporal variability of spectral power throughout the seizure, percentile-based summary statistics were computed across epochs. Specifically, the 5th, 10th, 25th, 50th, 75th, 90th and 95th percentiles were calculated for each of the seven frequency bands, resulting in 7 total and band-specific power representations × 7 percentile statistics = 49 absolute band power features. In addition to absolute power, relative band power features were computed to characterise the proportion of total spectral activity represented by each frequency band. For each frequency band, power was normalized by the total 1-30 Hz power prior to log-transformation. The same seven percentile statistics were subsequently computed across epochs, resulting in 6 relative frequency-band representations × 7 percentile statistics = 42 relative band power features. Together, the global band power feature set therefore consisted of 91 features describing both the absolute and relative spectral distribution of seizure activity across time.

#### Segment-Based Band Power Features

ECT seizures are inherently non-stationary, with spectral characteristics and morphology evolving throughout the recording. To capture these temporal dynamics, band power features were further quantified within predefined temporal segments. These segments were created using two complementary strategies. Absolute segmentation extracted four predefined time windows from each recording: the first five seconds, the middle ten seconds, the last ten seconds, and the final five seconds. Relative segmentation divided each recording according to its total duration, extracting the first 20%, the middle 40-60%, and the final 20% of the signal. For each segment, PSD was estimated over the segment signal, and a single log-transformed band power value was computed for each frequency band (total, delta, theta, low alpha, high alpha, low beta, and high beta). This resulted in 7 temporal segments × 7 frequency-band power representations = 49 segment-based power features.

#### Spectral Shape Features

In addition to band-specific power representations, three complementary features were computed to characterize the overall shape and distribution of the power spectrum across frequencies. These features were intended to capture broader spectral characteristics not fully represented by individual frequency-band power measures. PSD values were first averaged across the two EEG channels while preserving the epoch-wise structure. Each spectral shape feature was subsequently computed for individual epochs and averaged across epochs to obtain a robust global estimate for the recording.

The spectral centroid was computed as the power-weighted mean frequency:

$$f_{centroid} = \frac{\sum_f f \cdot P(f)}{\sum_f P(f)}$$

where *P(f)* denotes the power spectral density at frequency *f*. The spectral centroid reflects the center of mass of the spectrum and provides a measure of whether spectral power is concentrated at lower or higher frequencies.

To estimate the dominant frequency component of the seizure activity, the PSD was smoothed using a Gaussian filter (scipy.ndimage.gaussian_filter1d) prior to identifying the peak frequency, defined as the frequency corresponding to the maximum value of the smoothed spectrum.

Spectral entropy was computed from the normalized PSD as:

$$H = -\sum_f p(f) \cdot \log p(f) \text{ where } p(f) = \frac{P(f)}{\sum_f P(f)}$$

Spectral entropy quantifies the dispersion and complexity of spectral power across frequencies, where lower values indicate more concentrated spectral activity and higher values indicate a broader distribution of power across the spectrum. The spectral shape feature set therefore consisted of spectral centroid, peak frequency and spectral entropy.

**Temporal Dynamics**

In addition, the temporal evolution of seizure power was characterized using the total log-transformed power across epochs. Three features were computed: a linear trend in power over time, the difference in mean power between the first and second half of the recording, and the variance of power across epochs.

### Clinically Motivated EEG Features

In addition to spectral representations, features were extracted to reflect EEG characteristics commonly evaluated during routine clinical assessment of ECT seizures. According to clinical guidance from collaborating psychiatrists, signal amplitude, channel coherence and postictal suppression constitute important components of visual EEG-based seizure quality assessment during ECT. Consequently, amplitude-, suppression- and coherence- based features were included to quantitatively represent these clinically relevant signal characteristics. To capture both overall seizure characteristics and their temporal evolution throughout the recording, these features were computed both globally and within the predefined temporal segments.

### Amplitude Features

Amplitude was estimated using the analytic signal obtained via the Hilbert transform, implemented using hilbert from Scipy. For a signal x(t), the instantaneous amplitude (envelope) was computed as:

$$A(t) = |H(x(t))|$$

where H(·) denotes the Hilbert transform. This approach provides a smooth estimate of signal amplitude independent of oscillatory phase.

Amplitude features were computed both globally across the full recording and locally within each of the seven predefined temporal segments, using signals averaged across the two EEG channels. For the full recording and for each temporal segment, three statistical descriptors of the amplitude envelope were computed: mean amplitude, standard deviation of amplitude, and maximum amplitude. This resulted in 3 global amplitude features and 7 temporal segments × 3 amplitude descriptors = 21 segment-based amplitude features.

To further characterize temporal changes in seizure amplitude throughout the recording, two amplitude ratio features were additionally computed for the absolute segments: the ratio between middle-segment and start-segment mean amplitude, and the ratio between end-segment and middle-segment mean amplitude. These ratio features were included to capture temporal changes in seizure activity throughout the recording. Together, the amplitude feature set consisted of 3 global amplitude features, 21 segment-based amplitude features, and 2 temporal amplitude ratio features, resulting in a total of 26 amplitude-based features.

**Coherence Features**

Functional connectivity and synchronization between the two EEG channels were quantified using magnitude-squared coherence. These features were included because the degree of synchronization between EEG channels is considered an important aspect of seizure quality assessment in ECT.

For two signals *x* and *y*, coherence at frequency *f* is defined as:

$$C_{xy}(f) = \frac{|P_{xy}(f)|^2}{2 \cdot P_{xx}(f) \cdot P_{yy}(f)}$$

where $P_{xx}(f)$ and $P_{yy}(f)$ denote the power spectral densities of the individual signals and $P_{xy}(f)$ their cross-spectral density. Coherence values range from 0 to 1 and reflect the degree of linear correlation between the signals at each frequency. Coherence was estimated using Welch's method as implemented in scipy.signal.coherence, using 1-second windows (200 samples) with 50% overlap, consistent with the PSD estimation procedure.

Two complementary types of coherence features were extracted. First, coherence was computed separately within each of the seven predefined temporal segments of the seizure. For each segment, coherence values were averaged across the full 1-30 Hz frequency range, resulting in seven segment-specific coherence features that captured how synchronization changed over the course of the seizure. Second, band-specific coherence features were computed across the entire seizure recording. For each of the six non-total frequency bands, summary statistics of the coherence distribution were extracted, providing frequency-specific measures of synchronization between the EEG channels. For each band, coherence values were summarized using mean coherence, median coherence and 95th percentile coherence. This resulted in 6 frequency bands × 3 coherence summary statistics = 18 global band-specific coherence features. Together, the coherence feature set therefore consisted of 7 segment-level features and 18 global band-specific features, resulting in a total of 25 coherence-based features.

#### Suppression Features

Postictal suppression is an important aspect of EEG-based seizure quality assessment in ECT. Clinicians evaluate the extent to which EEG activity decreases following the seizure, as stronger suppression is generally associated with higher seizure quality. To quantitatively characterize suppression dynamics, suppression-related features were derived from the total log-transformed spectral power across epochs.

Three complementary suppression features were computed:

- Suppression range, defined as the difference between the 95th and 5th percentiles of log-transformed power
- Suppression interquartile range (IQR), defined as the difference between the 75th and 25th percentiles of log-transformed power
- Suppression ratio, defined as the ratio between the 95th and 5th percentile power values computed in the original power domain

These features were included as suppression-related summary measures, capturing the contrast between low- and high-power periods in the recording. The suppression feature set therefore consisted of 3 suppression-based features.

#### Clinical Variables and Treatment Features

In addition to the EEG-derived features, clinical variables were extracted from the structured clinical dataset and considered as potential predictors in the modelling framework. In this thesis, these variables were divided into two groups: clinical variables and anaesthesia-related variables. Clinical variables refer to demographic, treatment-related, and medication-derived variables, whereas anaesthesia-related variables refer specifically to the anaesthetic agents administered during the ECT procedure. The clinical variables used in the modelling framework were:

- patient sex,
- age at treatment,
- electrode placement,
- treatment number within series,
- seizure duration,
- motor seizure duration,
- stimulus dose, expressed as relative stimulus intensity corresponding to the percentage of the device's maximum output,
- cumulative benzodiazepine exposure, expressed as lorazepam-equivalent dose,
- cumulative antiepileptic drug exposure, expressed relative to defined daily dose.

These variables were selected because they describe patient characteristics, treatment context, stimulation parameters, and medication exposure that likely influences seizure characteristics and treatment response. The medication-derived variables are described in more detail below, while anaesthesia-related variables are treated as a separate feature group in the following subsection.

#### Medication Features

Medication-derived features were included because several medications can influence seizure threshold and EEG characteristics during ECT. Since medications within the same therapeutic class may differ in dose scale and pharmacological potency, doses were normalized before being used as model features.

#### Benzodiazepines

Benzodiazepine exposure was summarized as a cumulative lorazepam-equivalent dose. For each treatment session, up to five benzodiazepine administrations could be recorded, each with an Anatomical Therapeutic Chemical (ATC) code and administered dose. Recorded doses were converted to numeric values, standardized, and transformed into lorazepam-equivalent doses using ATC-specific conversion factors. The converted doses were then summed within each treatment session to obtain a single benzodiazepine exposure variable, expressed in lorazepam-equivalent milligrams.

#### Antiepileptic Drugs

Antiepileptic drug (AED) exposure was summarized as a cumulative dose normalized by World Health Organization defined daily dose (DDD) values. For each treatment session, up to three AED administrations could be recorded, each with an ATC code and administered dose. Each dose was divided by the corresponding ATC-specific DDD value, representing the dose relative to the typical daily therapeutic dose for that medication. The normalised doses were then summed within each treatment session to obtain a single cumulative AED exposure variable.

#### Anaesthesia Features

Additional features describing the anaesthesia administered during each treatment were derived. For each treatment session, up to seven anaesthesia administrations could be recorded, each containing an ATC code and a corresponding dose. Three anaesthetic agents commonly used during ECT were considered in the analysis: thiopental, propofol, and etomidate. For each treatment session, the total administered dose of each drug was calculated by summing the doses associated with the corresponding ATC codes which resulted in three new variables representing the total dose of thiopental, propofol, and etomidate administered during the procedure. If no anaesthesia ATC codes were recorded, or if the total administered dose across all anaesthetic agents was zero, the anaesthesia variables were treated as missing. Among treatments with valid labels, only 745 had at least one anaesthesia type and dose recorded. Consequently, models incorporating anaesthesia features were trained exclusively on this subset of 745 treatments.

#### Final Features

After feature extraction, the EEG-derived features were linked back to the clinical treatment records using the matched EEG filenames. The resulting dataset contained one observation per treatment session, with clinical variables, extracted EEG features and the expert relabelled seizure quality label. This dataset formed the basis for the subsequent modelling analysis.

**Table S1**: F1-score and Balanced Accuracy for all models in classification of observed seizure quality

| Model | Configuration | F1-score | Balanced Accuracy |
|---|---|---|---|
| Probit (Data Splits) | EEG, Weights | 0.665 ± 0.023 | 0.809 ± 0.022 |
| | EEG, No Weights | 0.605 ± 0.020 | 0.661 ± 0.019 |
| | EEG+Clin., Weights | 0.656 ± 0.014 | 0.788 ± 0.024 |
| | EEG+Clin., No Weights | 0.583 ± 0.036 | 0.650 ± 0.026 |
| Probit (Initialisation) | EEG, Weights | 0.646 ± 0.050 | 0.790 ± 0.026 |
| | EEG, No Weights | 0.610 ± 0.008 | 0.660 ± 0.009 |
| | EEG+Clin., Weights | 0.630 ± 0.028 | 0.740 ± 0.029 |
| | EEG+Clin., No Weights | 0.585 ± 0.022 | 0.643 ± 0.014 |
| OPANN-1 | EEG, Weights | 0.546 ± 0.055 | 0.732 ± 0.026 |
| | EEG, No Weights | 0.458 ± 0.047 | 0.528 ± 0.090 |
| | EEG+Clin., Weights | 0.547 ± 0.060 | 0.752 ± 0.027 |
| | EEG+Clin., No Weights | 0.504 ± 0.036 | 0.614 ± 0.029 |
| OPANN-2 | EEG, Weights | 0.592 ± 0.044 | 0.761 ± 0.035 |
| | EEG, No Weights | 0.481 ± 0.021 | 0.615 ± 0.028 |
| | EEG+Clin., Weights | 0.571 ± 0.044 | 0.749 ± 0.024 |
| | EEG+Clin., No Weights | 0.476 ± 0.009 | 0.619 ± 0.022 |
| Decision Tree | EEG, Weights | 0.703 ± 0.041 | 0.758 ± 0.025 |
| | EEG, No Weights | 0.708 ± 0.032 | 0.714 ± 0.032 |
| XGBoost | EEG, Weights | 0.792 ± 0.015 | 0.837 ± 0.016 |
| | EEG, No Weights | 0.780 ± 0.029 | 0.741 ± 0.032 |

**Table S2**: Binary classification performance in classification of observed seizure quality

| Model | F1-score | Balanced Accuracy | AUC (T vs A) |
|---|---|---|---|
| Probit | 0.770 ± 0.020 | 0.841 ± 0.019 | 0.925 ± 0.009 |
| OPANN-1 | 0.762 ± 0.010 | 0.880 ± 0.008 | 0.948 ± 0.006 |
| OPANN-2 | 0.761 ± 0.008 | 0.877 ± 0.012 | 0.946 ± 0.006 |
| Decision Tree | 0.770 ± 0.011 | 0.834 ± 0.014 | 0.844 ± 0.016 |
| XGBoost | 0.829 ± 0.007 | 0.861 ± 0.006 | 0.930 ± 0.014 |

**Table S3**: Multiclass predictive performance in prediction of subsequent seizure quality

| Model | F1-score | Balanced Accuracy | AUC (0 vs T) | AUC (T vs A) |
|---|---|---|---|---|
| Linear Probit | 0.3485 ± 0.0198 | 0.4376 ± 0.0475 | 0.5533 ± 0.0938 | 0.6428 ± 0.0471 |
| OPANN-2 | 0.3115 ± 0.0099 | 0.3398 ± 0.0275 | 0.5632 ± 0.0698 | 0.5470 ± 0.0474 |
| Decision Tree | 0.3726 ± 0.0174 | 0.4339 ± 0.0284 | 0.4015 ± 0.0832 | 0.6314 ± 0.0517 |
| XGBoost | 0.3911 ± 0.0117 | 0.4300 ± 0.0249 | 0.4312 ± 0.0513 | 0.7118 ± 0.0273 |
